# Mobility and ART retention among men in Malawi: a mixed methods study

**DOI:** 10.1101/2022.11.07.22281938

**Authors:** Marguerite Thorp, MacDaphton Bellos, Tijana Temelkovska, Misheck Mphande, Morna Cornell, Julie Hubbard, Augustine Choko, Tom Coates, Risa Hoffman, Kathryn Dovel

**Author notes:** Corresponding author: Marguerite Thorp, 10833 Le Conte Ave., 52-215 CHS, Los Angeles, CA 90095-1688. E-mail addresses of authors.

## Abstract

**Introduction:** Mobility is associated with worse outcomes across the HIV treatment cascade, especially among men. However, little is known about the mechanisms that link mobility and poor HIV outcomes and what types of mobility most increase the risk of treatment interruption among men in southern Africa.

**Methods:** From August 2021 – January 2022, we conducted a mixed-methods study with men living with HIV (MLHIV) but not currently receiving antiretroviral therapy (ART) in Malawi. Data collection was embedded within two larger trials (ENGAGE and IDEaL trials). We analyzed baseline survey data of 223 men enrolled in the trials who reported being mobile (defined as spending ≥14 nights away from home in the past 12 months) using descriptive statistics and logistic regression. We then recruited 32 men for in-depth interviews regarding their travel experiences and ART utilization. We analyzed qualitative data using constant comparative methods.

**Results:** Survey data showed that 34% of men with treatment interruptions were mobile, with a median of 60 nights away from home in the past 12 months; 69% of trips were for income generation. More nights away from home in the past 12 months and having fewer household assets were associated with longer periods out of care. In interviews, men reported that travel was often unplanned, and men were highly vulnerable to exploitive employer demands, which led to missed appointments and ART interruption. Men made major efforts to stay in care but were often unable to access care on short notice, were denied ART refills at non-home facilities, and/or were treated poorly by providers, creating substantial barriers to remaining in and returning to care. Men desired additional multi-month dispensing (MMD), the ability to refill treatment at any facility in Malawi, and streamlined pre-travel refills at home facilities.

**Conclusions:** Men prioritize ART and struggle with the trade-offs between their own health and providing for their families. Mobility is an essential livelihood strategy for MLHIV in Malawi, but it creates conflict with ART retention, largely due to inflexible health systems. Targeted counseling and peer support, access to ART services anywhere in country, and MMD may improve outcomes for mobile men.

## 1. Introduction

Antiretroviral therapy (ART) treatment interruptions are common throughout sub-Saharan Africa (SSA), particularly for men(1-4). Treatment interruptions contribute to poor health outcomes, antiretroviral drug resistance, and further HIV transmission(5-7). Travel is a frequent reason for missing ART appointments and can lead to extended treatment interruptions due to numerous barriers to returning to care(8, 9). Extended treatment interruptions due to mobility may explain the poor rates of ART retention and viral suppression experienced by mobile people throughout SSA(10-16).

Mobility in SSA is common and is not new(17-19). In a 2005 survey conducted in Malawi, 25% of men aged 25-49 years had permanently moved to a new village within the last five years(19). This figure does not include temporary mobility, which is likely even more common(20). Throughout the region, people engage in temporary mobility for many reasons, including work, trade, cultural or religious events, and funerals. However, temporary mobility is rarely captured in mobility data and to date has been understudied.

Men living with HIV (MLHIV) are as likely to be mobile as those without HIV, and some evidence suggests they are more mobile(21, 22). There is an association between mobility and HIV infection, but the causal direction is not clear: mobile people might be at higher risk of infection, or MLHIV may have greater need for mobility, whether due to discrimination experienced at home, marital dissolution, or the need to recover from financial losses experienced during prolonged illness(23). Mobility (including temporary mobility) among MLHIV deserves more attention because it is associated with poor HIV outcomes and is likely to increase in the coming decade(24, 25).

Most research has treated mobility as a uniform phenomenon, with single metrics defining “mobile” and “non-mobile” populations obscuring the heterogeneity of travel experiences(26). However, different aspects of mobility – who travels where, why, for how long, and with whom – may affect men’s use of ART services differently(27). To our knowledge, only one study has explored the impact of varying forms of mobility on ART use. The study found that longer trips (averaging more than two nights) among ART clients in Kenya and Uganda were associated with significantly lower levels of treatment adherence as compared to shorter trips (averaging one to two nights)(28). Additional research is needed to assess whether this relationship exists in other settings, if other aspects of mobility such as destination and purpose also differentially impact use of ART services, and how forms of mobility affect facility attendance for routine ART refill appointments.

We performed a mixed-methods study among MLHIV with a history of ART interruption in Malawi to characterize different forms of mobility and how mobility influenced treatment engagement.

## 2. Methods

We conducted a sequential mixed methods study including surveys and in-depth interviews with MLHIV in Malawi. We analyzed data from two parent trials enrolling MLHIV who experienced treatment interruption to understand the prevalence and forms of mobility among this population. We then selected a subset of mobile men for in-depth interviews (IDIs) exploring how mobility influenced men’s engagement in care and what men did to overcome mobility-related barriers to care.

### 2.1 Trial context

Identifying Efficient Linkage Strategies for Men in Malawi (“IDEaL,” BMGF INV-001423) and Engaging Men Through Differentiated Care to Improve ART Initiation and Viral Suppression (“ENGAGE,” NIH R01 MH122308-02) are two trials are being conducted in 24 high-burden health facilities across eight districts in Malawi, a country with an HIV prevalence of 8.1%(29) and a national HIV program offering free ART to all PLHIV. Detailed protocols of the parent trials can be found elsewhere(30). Briefly, the trials test four interventions to improve men’s initiation and re-initiation and six-month retention in HIV care: male-specific counseling, home-based ART services (for one and three months), and ongoing motivational interviewing. Eligibility criteria for both parent trials are: (1) being male and ≥15 years of age, (2) living in facility catchment area, and (3) tested positive for HIV but did not initiate ART (within seven days of positive test) or initiated ART but currently experiencing treatment interruption. Treatment interruption was defined as either (a) initiated ART but missed the first 30-day refill appointment by seven days or more, or (b) initiated ART and attended first refill but was ≥28 days late for a subsequent refill appointment. We identified potentially eligible men through medical chart reviews at participating facilities and then traced them at home for study screening and, if eligible, enrollment. This sub-study was embedded in the baseline data collection for both parent trials. Trial enrollment is complete but follow-up data collection is still ongoing.

### 2.2 Survey data collection & analysis

#### Data Collection

Our sample of MLHIV were enrolled in the parent trials between August 25, 2021 and January 5, 2022. Within the parent trials, all men completed baseline surveys immediately following enrollment. Survey domains included socio-demographics, ART treatment history, travel history within the past 12 months, and anticipated travel in the next 12 months. Men who previously initiated ART were asked the date of the refill appointment that they last missed, and study staff verified data using medical chart reviews at participating health facilities.

#### Variable Definitions

For this sub-study, we defined mobility as spending ≥14 nights away from home in the past 12 months. We included several measures to explore forms of mobility, including total nights spent away from home in the past 12 months, number of trips with ≥14 consecutive nights away from home within 12 months, and details about each trip ≥14 consecutive nights including destination, length, and purpose. Our primary outcome of interest was total number of days out of ART care (either because an individual never initiated ART or had treatment interruption from routine ART refills). We calculated days out of care as the number of days between the most recent missed appointment and the date of study enrollment. We included other descriptive variables about men’s use of HIV services, including years since HIV diagnosis and ever initiated ART.

We included the following sociodemographic variables: age, marital status (monogamous or polygamous vs. single or divorced/widowed), having financial savings (yes/no), and received any secondary school education (yes/no). To measure wealth we used a 15-item household asset index used widely in the region(31). We used the first dimension of a principal component analysis, linearly transformed to a scale of 0 to 10 to make the scores more easily interpretable. Having access to a cell phone is included in the asset index but also analyzed separately given its importance for communication while traveling.

#### Analysis of quantitative data

We used descriptive statistics to explore forms of mobility and socio-demographic characteristics. Men could report more than one trip, allowing us to analyze incidence of mobility forms and heterogeneity across time. We used a chi-squared test to compare destinations of income-generating and non-income-generating trips. We conducted univariable truncated negative binomial regressions to explore associations between mobility forms and days out of care. Any variable that was associated with a p-value of <0.10 in a univariable regression was included in a multivariable truncated negative binomial regression model. We conducted all analyses in Stata (17.0, Stata Corp LLC, College Station, TX).

### 2.3 In-depth interview data collection & analysis

#### Data Collection

We purposively sampled men who self-reported being mobile during baseline surveys for IDIs to achieve balanced representation of region (North, Central, South) and travel destination (domestic or international). A member of the study team (MB) contacted selected men via phone or home visit if phone was not available. All interviews took place in the community at a location and time of the participant’s choice.

#### Interview Guide

The interview guide used a narrative approach(32) to elicit information about recent travel, experiences with ART, and how travel affected men’s ability to continue ART. Men were asked to describe their most recent trip, their longest trip in the last year, any travel experiences that caused them to miss ART refill appointments or run out of medication, and recommendations for how ART services could better serve them and other mobile men. The guide was piloted with seven participants and revised based on feedback and review of transcripts. Interviews were conducted in a local language (Chichewa) and audio recorded, transcribed, and translated into English.

#### Analysis of qualitative data

Three study team members (MT, TT, KD) developed a codebook based on key concepts from the interview guide and emergent codes from a review of the first seven interviews. Interviews were coded using Atlas.ti Mac (Version 9.1.2)(33). MT and TT piloted the codebook on two transcripts, resolved any disagreements with coding, and revised the codebook accordingly. The remaining transcripts were coded by either MT or TT. We wrote brief, reflective memos for each transcript to summarize key themes at the time of coding and used these to guide analysis(34). We analyzed coded text using constant comparative methods(35), comparing and contrasting themes by several forms of mobility: length; international vs. national travel; and planned vs. unplanned travel. Co-authors reviewed in detail summaries of each theme to develop qualitative results.

### 2.3 Ethical considerations

The research was approved by the Malawi National Health Sciences Research Committee (#20/0712567) and the UCLA IRB (#21-000209). Written consent was obtained from all respondents at the time of enrollment in the parent trials and additional oral consent was obtained for in-depth interviews.

## 3. Results

### 3.1 Participant characteristics

#### 3.1.1 Survey participants

Between August 2020 - January 2021, 651 men living with HIV but not actively in HIV care were enrolled in the parent trials. A total of 223/651 (34%) met the definition for “mobile” and were included in this sub-study (Figure 1).

**Figure 1.**
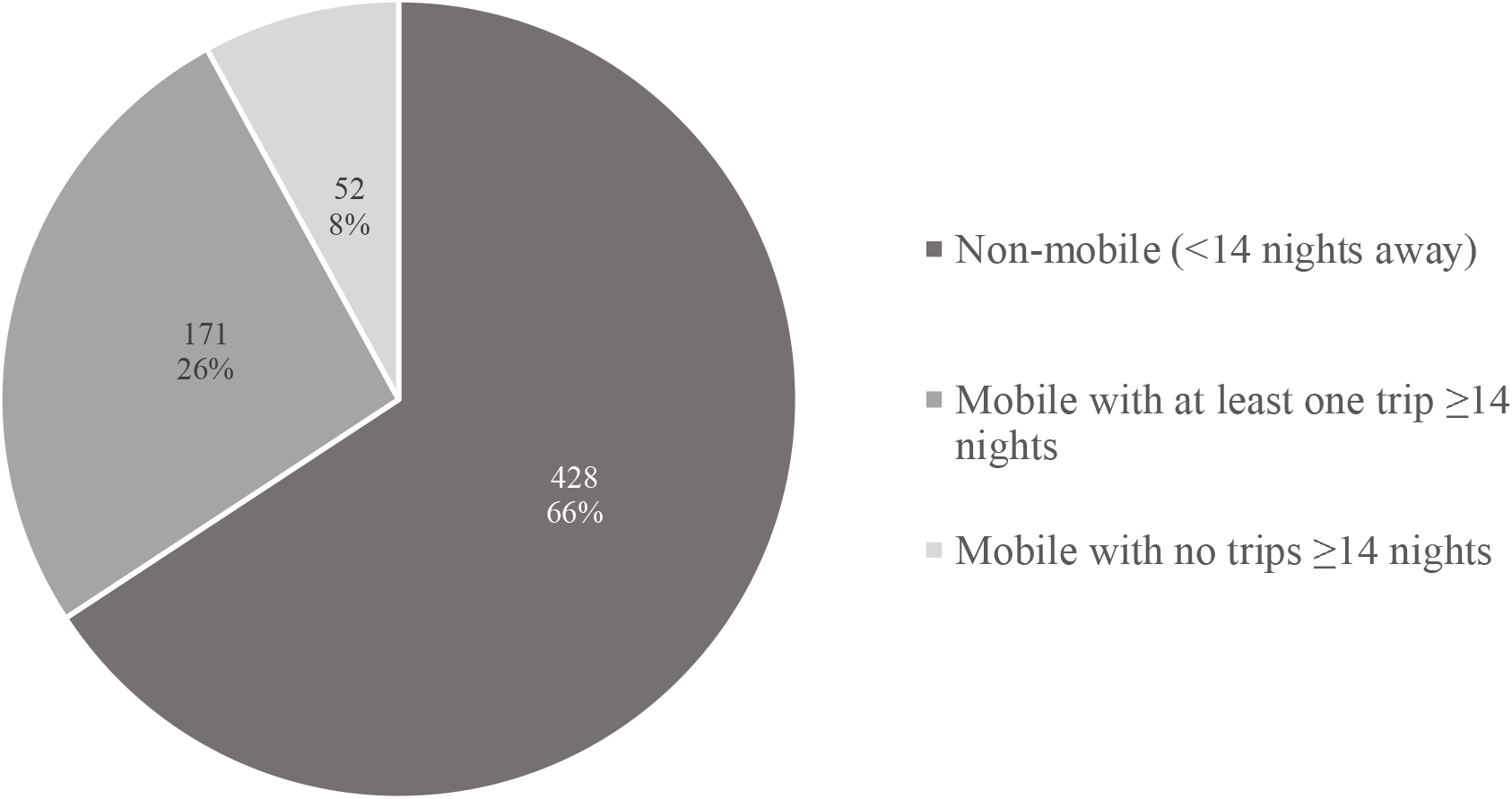
Men living with HIV disengaged from ART in Malawi. Proportion of participants in parent trials reporting mobility, including any long trips (≥14 nights) or only short trips (<14 nights).

Mobile men had a median age of 38 years (IQR:32-45) and were predominantly married (69%) (Table 1). They had a median of 3.2 years since HIV diagnosis; almost all men (90%) had previously initiated ART and were experiencing treatment interruption, while 22 (10%) had never initiated ART. Those with treatment interruption were out of care for a median of 78 days (IQR:48-190) prior to enrolling in the parent trials. The majority (197 [88%]) had disclosed their HIV status to someone. Nearly two-thirds of participants had access to a mobile phone in their household, 33% had some form of financial savings, and 68 (30%) had attended secondary school.

**Table 1.** Socio-demographics, ART history, and travel history of mobile men experiencing HIV treatment interruption in Malawi (n=223)

| **Demographics** |  |
| --- | --- |
| Age, median years (IQR) | 38 (32-45) |
| Married, n (%) | 153 (69%) |
| **ART Experience** |  |
| Years since diagnosis, median (IQR) | 3.2 (0.9-9.4) |
| Ever initiated ART, n (%) | 201 (90%) |
| Days out of care at enrollment, median (IQR) | 78 (48-90) |
| Disclosed HIV status to someone, n (%) | 197 (88%) |
| **Socioeconomic status** |  |
| Household access to a cell phone, n (%) | 136 (61%) |
| Has savings, n (%) | 74 (33%) |
| Attended any secondary school, n (%) | 68 (30%) |
| **Travel experience & plans** |  |
| Nights away from home in last year, median (IQR) | 60 (30-90) |
| Average trip length >30 days, n (%) | 93 (42%) |
| Took long trip ($\geq 14$ nights) in last year, n (%) | 171 (77%) |
| Planning travel ($\geq 14$ nights total) in next year, n (%) | 57 (26%) |
| Planning long trip ($\geq 14$ nights) in next year, n (%) | 46 (21%) |
| Number of trips $\geq 14$ nights planned in next year, n (%) | |
| 1 | 35 (16%) |
| 2 | 11 (5%) |

Mobile men spent a median of 60 nights away from home in the last 12 months, and nearly half the trips were on average >30 nights long. Over 75% had at least one long trip (≥14 consecutive nights) in the previous 12 months. Men with at least one long trip spent more nights away from home versus those with only short trips (<14 nights): 30 (IQR:18-60) versus 60 (IQR:30-90) median nights, respectively (not shown). Notably, of men who had been mobile in the previous year, only one quarter anticipated being mobile in the coming year, representing the unpredictable nature of travel.

Overall, 171 men reported 257 long trips (≥14 nights) (Table 2). Long trips lasted a median of 30 (IQR:14-90) nights. The majority (69%) of travel was for income generation, including work for wages or trading goods. Most long trips (68%) were to destinations within Malawi, predominantly within-region (53%). International travel comprised 32% of long trips and was almost exclusively to the neighboring countries of Mozambique and Tanzania. About half of the long trips were taken alone. Trips for income generation were more likely to be international than non-income generating trips.

**Table 2.**
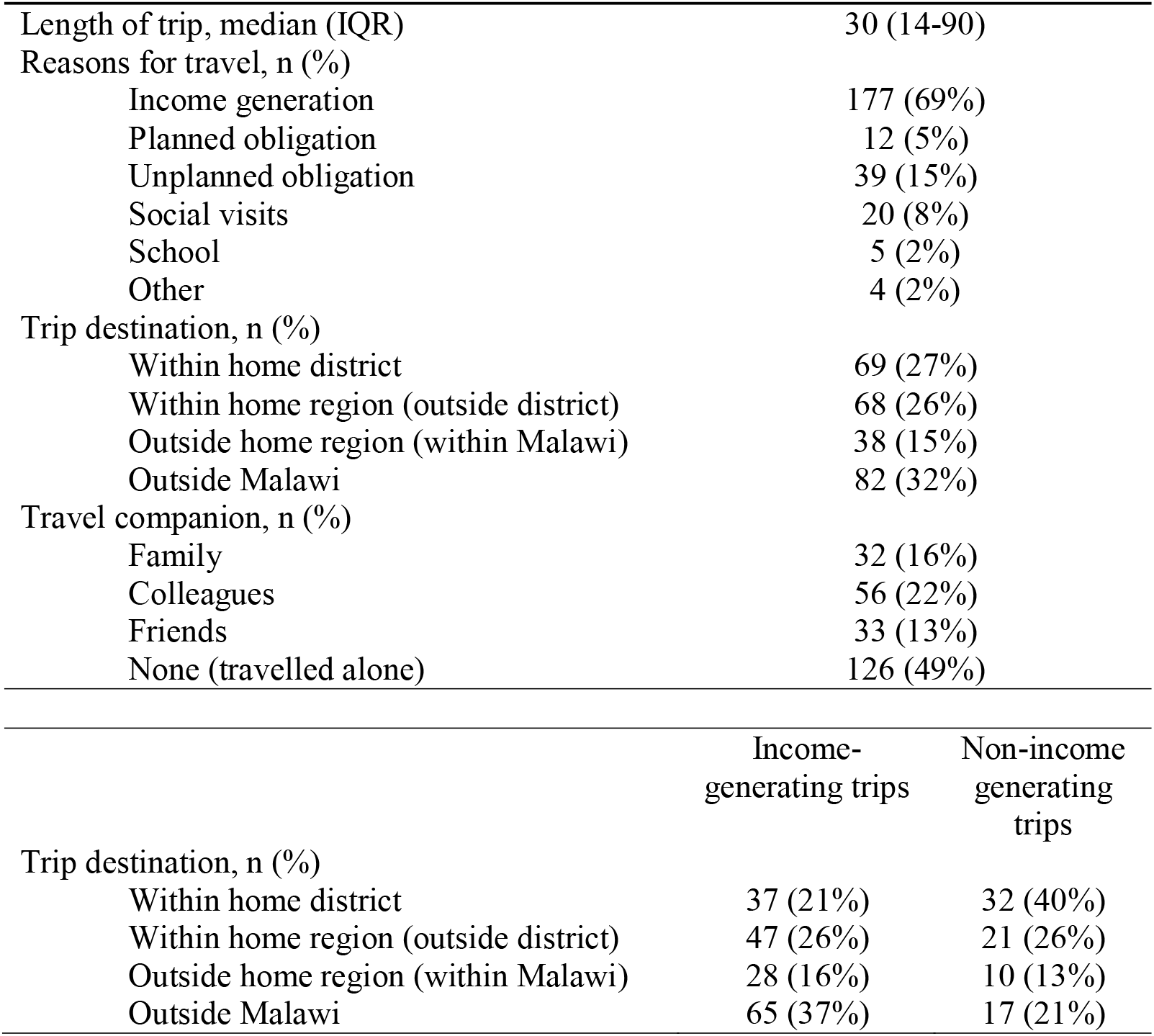
Trips ≥14 nights taken by mobile men experiencing treatment interruption in Malawi (n=257)

|  |  |
| --- | --- |
| Length of trip, median (IQR) | 30 (14-90) |
| Reasons for travel, n (%) |  |
| Income generation | 177 (69%) |
| Planned obligation | 12 (5%) |
| Unplanned obligation | 39 (15%) |
| Social visits | 20 (8%) |
| School | 5 (2%) |
| Other | 4 (2%) |
| Trip destination, n (%) |  |
| Within home district | 69 (27%) |
| Within home region (outside district) | 68 (26%) |
| Outside home region (within Malawi) | 38 (15%) |
| Outside Malawi | 82 (32%) |
| Travel companion, n (%) |  |
| Family | 32 (16%) |
| Colleagues | 56 (22%) |
| Friends | 33 (13%) |
| None (travelled alone) | 126 (49%) |

|  | Income-generating trips | Non-income<br>generating<br>trips |
| --- | --- | --- |
| Trip destination, n (%) |  |  |
| Within home district | 37 (21%) | 32 (40%) |
| Within home region (outside district) | 47 (26%) | 21 (26%) |
| Outside home region (within Malawi) | 28 (16%) | 10 (13%) |
| Outside Malawi | 65 (37%) | 17 (21%) |

Table 3 describes associations between mobility and days out of care. Of the 201 mobile men who had previously initiated ART, complete data for analysis of factors associated with treatment interruption was available for 195. In multivariable models, each additional month away from home in the previous 12 months was associated with a 9% increase in the length of treatment interruption (aRR 1.09, 95%CI 1.03-1.15). Men who had more assets were likely to experience shorter treatment interruption than those without assets (aRR0.87, 95%CI 0.87-0.92).

**Table 3.**
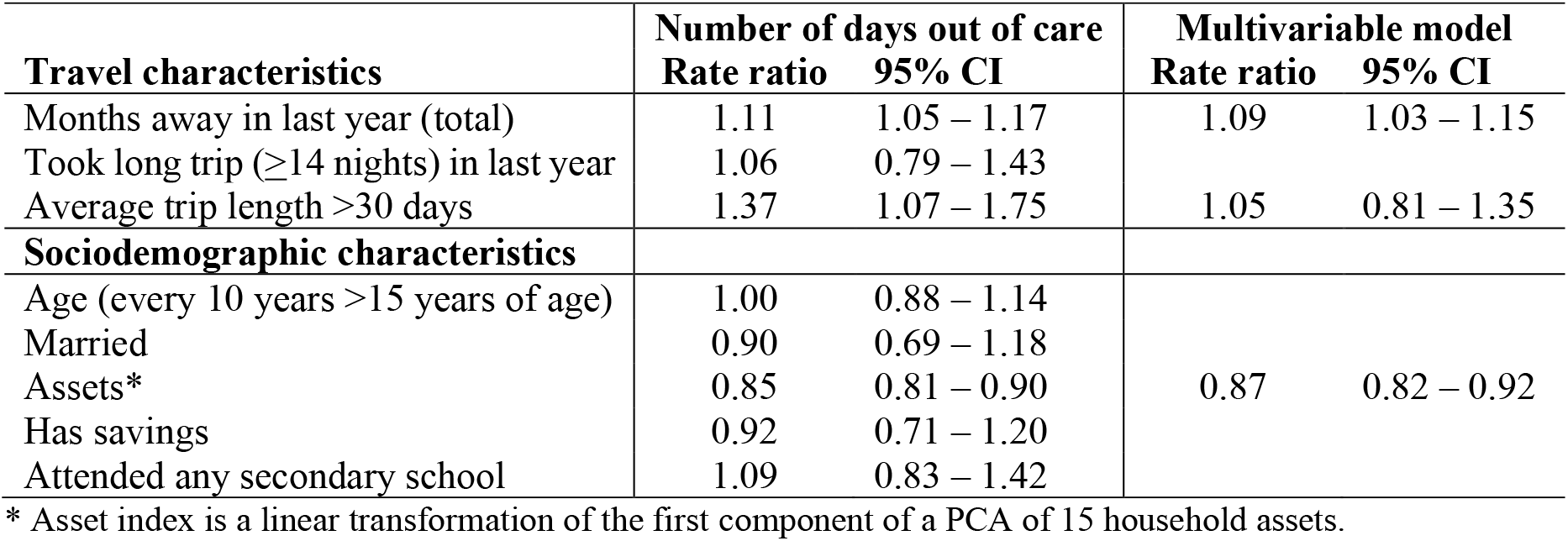
Factors associated with length of treatment interruption among mobile men in Malawi (n=195)

Table 3 describes associations between mobility and days out of care. Of the 201 mobile men who had previously initiated ART, complete data for analysis of factors associated with treatment interruption was available for 195. In multivariable models, each additional month away from home in the previous 12 months was associated with a 9% increase in the length of treatment interruption (aRR 1.09, 95%CI 1.03-1.15). Men who had more assets were likely to experience shorter treatment interruption than those without assets (aRR0.87, 95%CI 0.87-0.92).

Table 3 describes associations between mobility and days out of care. Of the 201 mobile men who had previously initiated ART, complete data for analysis of factors associated with treatment interruption was available for 195. In multivariable models, each additional month away from home in the previous 12 months was associated with a 9% increase in the length of treatment interruption (aRR 1.09, 95%CI 1.03-1.15). Men who had more assets were likely to experience shorter treatment interruption than those without assets (aRR0.87, 95%CI 0.87-0.92).

#### 3.1.2 In-depth interview participants

Between August 5-December 2, 2021, we selected 39 men from six districts in Malawi for IDIs and completed IDIs with 32 men. We excluded seven men who were not traced or did not meet the mobility criteria. The median age was 39 (IQR:33-46) years and 6% had never initiated ART (Appendix A). The mean time since HIV diagnosis was 1.7 years (IQR:0.8-9.0). Overall, 27 (84%) were married and 13 (41%) had attended secondary school. In total, interviewees discussed 65 recent trips with a median length of 21 nights (IQR:7-75). Most travel was within Malawi; half of the men traveled to Mozambique or Tanzania at some point.

### 3.2 Characteristics of travel

#### 3.2.1 Reasons for travel

More than three-quarters of participants described traveling for income generation, usually essential for basic survival. Forms of income generation varied widely and included work in agriculture, construction, and trading goods. Most work was *ganyu*, or “piece work,” in which men were hired for a specific task or number of days with no formal contract, worker protection, or job security. A few men reported unique jobs such as traditional healing, traditional dancing, and singing. Most men saw traveling as their only option for earning money.

> “Because of weather problems, sometimes you harvest, sometimes not – so that is why we plan a sudden trip unexpectedly, because you see that there is nothing at home. But if we have enough maize, we settle [i.e. stay at home].” *Age 40-49, 120 nights away, traveled for piece work*

Several participants described inhumane working conditions or not being paid what they were promised. Men had little information about when they would travel, frequently had only one or two days to prepare for trips that could last months, and the duration was often unknown when departing.

> “I was unprepared for this trip. A person just found us [a group of men] and said he wanted men for a job, so we should go to Mzuzu… so I just left, and I did not even bid anyone farewell at home. [My family] just heard about [the trip] when I was already there.” *Age 20-29, 300 nights away, traveled for piece work*

Other men traveled without a plan for employment, hoping to find work when they arrived at their destination.

> “I usually travel to Mozambique to look for some piece jobs, like farming. In return they give me maize and [I] come back home to feed the family. I travel on foot to and from. It’s about a 20km journey.… We usually don’t count the days when we go work. Sometimes we stay longer, until I achieve what I went for. So I don’t really have a timeline. If there is less work, I stay for less days, and stay more days if there is more work.” *Age 40-49, 20 nights away, traveled for piece work*

Other travel occurred in response to urgent life events, including caregiving for ill relatives, funerals, or domestic disagreements. Caregiving trips were unexpected and often lasted longer than intended as illnesses progressed and sometimes led to death, with subsequent funeral obligations.

> “On this trip, I left suddenly when my [relative] passed away in Mchinji. I stayed there for a month.… If I [suggested] leaving to return back home, [my family] would stop me and say it has been too short a time.” *Age 30-39, 150 nights away, traveled for funeral*

#### 3.2.2 Planned vs. unplanned travel

Men were largely unable to predict or plan their travel. Some travel was inherently unpredictable, such as travel for caregiving, funerals, or domestic quarrels. Work travel was also frequently unplanned because of men’s vulnerable economic circumstances, obliging them to take any available income opportunities. Men were often recruited for piece work the day before departure; formal employment was rare, but even employed men frequently received minimal notice from their employer.

> “Most of the times when you are doing piece jobs you don’t have complete freedom to travel when you want. You [rely on] the person who helped you get the job, you wait for them to say, ‘my work is over, let’s go.’” *Age 40-49, 127 nights away, traveled for piece work*
>
> “My main problem is that I have emergency trips like the boss will just call me and tell me that he has sent money for me to travel today, while the medicine [ART] is running out tomorrow or the day after that. If I refuse, that he should wait for me to organise a few things, it would be that he has already made all the arrangements [and hired someone else].” *Age 40-49, 270 nights away, traveled for piece work*

Men were frequently dependent on employers for travel money, with little autonomy over when they returned home. Transport challenges, especially the lack of bus fare, often delayed returns.

### 3.3 Mobility & HIV Care

#### 3.3.1 Treatment Interruption

All men interviewed reported valuing ART and recognizing that treatment was central to meeting their life goals. The majority of men brought their ART on trips, but most did not have enough for the full duration.

Importantly, trips with known duration and adequate preparation time never resulted in treatment interruptions. Most unplanned trips that resulted in medications running out lasted ≥30 nights. Short trips, even if unplanned, rarely led to treatment interruption.

> “I was expecting to only stay for five days and come back, so I didn’t take my health passport [personal medical record]. I did [bring ART]. But I took few pills because I wasn’t expecting to stay for a month…. They finished midway [through the trip]. It was hard for me because I didn’t have a health passport for me to go to the hospital.” *Age 30-39, 30 nights away, traveled for formal employment*

#### 3.3.2 Strategies to maintain adherence and engagement

Mobile men used various strategies to stay on ART while traveling. One common strategy was caregiver refills, whereby wives or siblings at home collected ART refills. However, it was still difficult to transport medication to men who were traveling. Men either needed to return home shortly after the caregiver refill appointment or make complex arrangements to have medications delivered.

> “My elder sibling [attended the ART refill appointment]. After they took them [ART], they managed to send them, and the pills reached me. They sent via minibus, they paid the conductor, we communicated via phone, and the pills got to me.” *Age 30-39, 30 nights away, traveled for piece work*

Other men tried to access emergency ART refills, whereby ART clients can obtain a one-month supply of ART at any Malawi health facility. Emergency refills are part of national protocol(36), but several men who attempted this process were denied treatment and told to return home to obtain a formal transfer letter.

> “When I went to [my family’s home], and went to the hospital there, they told me that I should get a transfer letter…. They did not give me [ART].” *Age 30-39, total nights away unavailable, traveled due to conflict with spouse*

One-third of participants returned home from travel specifically for ART refills; a few reported forfeiting gainful employment to obtain medication refills.

> “In Dar-es-Salaam, I was doing piece work, lifting bags for people.… I [brought ART], but they finished midway. That’s why I came back. I stayed three days without ARVs then I came back home.” *Age 20-29, 115 nights away, traveled for piece work*

A few men who had time to plan their travel obtained multiple months of medication prior to traveling so their ART supply could last until they returned home.

#### 3.3.3 Consequences of treatment interruption

Over half of the men who ran out of medication while traveling reported periods longer than one month out of care. They frequently reported feeling physically ill, with symptoms including generalized weakness, skin rashes, and weight loss. For many, feeling ill triggered their return from travel and re-engagement in HIV care.

> “I was worried because when I have the pills, I am stronger and do not get sick as often. When I ran out of the pills, I saw that I was sick, got malaria frequently and was just weak most of the time. That was when I saw that I could die, so I should just come back [to my home facility].” *Age 30-39, total nights away unavailable, traveled for piece work*

Treatment interruptions due to mobility also led to conflicts with healthcare workers (HCWs). Half of the men returning to care after travel-related treatment interruption reported poor treatment, such as shouting, scolding, or “punishment,” usually being made to wait and be seen last at the ART clinic.

> “It was an abusive system of welcoming… Like [HCW] asking why I was late… ‘you are not taking ARVs, what is happening?’ Those words hurt a lot.” *Age 40-49, 30 nights away, traveled to trade goods*
>
> “When you skip days, they call you a defaulter. Even if you go as early as you can, you are served last…. It’s hard on us. You go early in the morning, spend the whole day there, hungry, and also walk a long distance going back home.” *Age 50-59, total nights away unavailable, traveled for caregiving*

Men responded differently to negative HCW interactions: most men expressed understanding of the HCW’s behavior, but one third of men were upset by punitive treatment. Several men avoided re-engaging after travel because they feared a negative response from HCWs.

> “After I came [home] I had the idea to go to the hospital. At that time I was still feeling sick… but I was scared to go to the hospital because I didn’t know what happens in such cases. The ARVs had finished 19 days prior, so what would they [HCW] say? Won’t they shout at me? Right? I had worries and I just stayed [at home].” *Age 20-29, 300 nights away, traveled for piece work*

#### 3.3.4 Participant recommendations for improving retention on ART while mobile

Men suggested several ways to improve access to HIV care while traveling, including greater flexibility in when, where, and how often they refilled their ART.

Men desired easier pre-travel refills, with ART available any day of the week.

> “It should be that when you have to travel and you have five days more before you have to collect your ARVs, you can meet health workers and ask them to help you. Tell them you are traveling, but the date that they gave you [for your next refill] is very soon.” *Age 40-49, 60 nights away, traveled for piece work and caregiving*

Men also wanted emergency refills to be available nationwide so they could refill their script anywhere in Malawi without requiring a transfer letter. One man suggested providing a list of facilities where ART would be available while travelling.

> “They need to make a list [of facilities] for the men so that they can continue taking ARVs wherever they are going…. If I travel to Lilongwe, I can go to other hospitals and explain that I am travelling [and get ART].” *Age 20-29, 20 nights away, traveled to receive traditional healing*

Finally, several men suggested providing multi-month dispensing to prevent missing appointments. Few men knew that multi-month dispensing was already standard policy in Malawi.

> “We should come to the hospital to report [travel]… but the time we will stay there [traveling] is unknown, so they should give us more medicine.… If they [usually] give us one bottle [of ART], they should give us maybe two because it is unknown when we will return.” *Age 30-39, 30 nights away, traveled for formal employment*

## 4. Discussion

In this mixed-methods study in Malawi, we found high rates of mobility among MLHIV who experienced treatment interruption (34%), with men spending on average 60 nights away from home in the previous year. Mobile men traveled often and most took trips longer than 14 nights. Men who spent more total time away experienced more days outside HIV care but having household assets was protective against treatment interruption. Mobility was essential for income generation for MLHIV, but created unavoidable conflicts with ART appointments, especially when travel was unplanned. In contrast to prevailing narratives about men, we found that men valued their HIV care and made efforts to remain on treatment despite unplanned travel. Returning to care was often difficult due to ongoing travel, rigid health systems that denied emergency and caregiver refills, and fear of punitive treatment from HCWs. Mobile men desired system-level adaptations such as multi-month dispensing, easier pre-travel refills, and emergency refills at facilities in their destination.

Mobility was a critical livelihood strategy for our study participants, as is the case in general populations throughout SSA(24, 37). Participants in our study sought to support their families and live up to the masculine ideal of a provider, but they were impeded by the realities of poverty(38). They stressed that travel for work was not their choice, but a last resort in desperate times. Malawi is one of the poorest countries in the world(39). In rural Malawi, subsistence farming dominates but is often inadequate to meet a family’s needs, resulting in steep competition for few cash-earning opportunities(40, 41). Men working in *ganyu* arrangements to feed themselves and their families must be willing to travel at a moment’s notice to pursue any available income. Consistent with studies showing *ganyu* workers’ weak bargaining power(42), men described many forms of vulnerability in their work arrangements, including physically difficult conditions, not being paid what they were promised, working for longer than expected, and dependence on employers for food, housing, and transportation funds. Even if men wanted to leave early to refill their medications, they often lacked funds for return travel. Aside from work, men also traveled for family obligations including attending funerals and caregiving, essential activities in a setting where family ties are often the only available insurance against severe deprivation(43, 44). Men faced challenging trade-offs between their own well-being (including staying on ART) and their desire to provide for their families and fulfill social expectations.

In line with a growing body of research on men’s participation in HIV care(45), we found that men valued their HIV treatment, were aware of the consequences of treatment interruption(46), and made substantial efforts to stay in care(47). Men modified their travel plans when possible and utilized the few options available within the health system, such as unscheduled refills before traveling and emergency refills at health facilities in their destination. Our findings conflict with a prevailing narrative framing men as difficult clients, unwilling to seek HIV care, and affirm the growing recognition that men are and should be treated as active participants in their own healthcare(48).

Despite men’s expressed desire to stay in treatment, adherence strategies were often rendered ineffective by a rigid health system. Emergency and caregiver refills were denied by cautious HCWs. Limited ART clinic days prevented pre-travel refills when travel was unplanned. Since Malawi adopted “Test & Treat” in 2016, there has been increasing support for male-friendly care interventions(49), and national guidelines were modified in February 2022 to allow up to three-month refills for individuals re-initiating care. However, though these changes may benefit mobile men indirectly, there has been almost no evolution of the policies on mobility. In the current era of dolutegravir-based regimens – in which resistance risk is low and ART supply chains operate smoothly – there is an opportunity to pursue client-centered care in ways that were previously considered too risky.

Mobile men identified several solutions to their challenges, including the ability to obtain refills at any ART site in Malawi. A national electronic health record would allow fully portable care, making this strategy more feasible. Mobile clients – along with all clients – would also benefit from a shift in health system culture to a non-paternalistic approach, in which clients are trusted to refill at the most convenient location. More immediate action could also be taken by expanding access to emergency refills and multi-month dispensing (MMD). Men in our sample commonly reported running out of medications on long trips, and a one-month refill without easy access to emergency refills is inadequate. Emergency refills are officially a health system policy, and six-month refill periods are highly effective for stable clients in Malawi(50-52), but most men in our study had only received monthly or three-month refills. This may be due to limited scale-up of the six-month MMD policy(50) or because participants were not regarded as virologically stable, though they would have benefited greatly from MMD. A comprehensive solution to minimize treatment interruption for mobile men would include MMD, streamlined pre-travel refills, more accessible emergency refills, and peer support to increase awareness of existing options and help men access these.

This study has several limitations. We only included men who experienced treatment interruption, but some mobile MLHIV are able to remain on ART, and they or the nature of their travel may differ from study participants. Results may be influenced by social desirability bias if men over-reported their desire to remain on ART and their efforts to do so. Findings may be subject to recall bias, as some treatment interruptions took place more than one year prior to the interview. Finally, we were surprised to see little travel to South Africa given Malawi’s long-standing role as a labor reserve for South Africa(53, 54). This may be due to reduced migration due to COVID-19(55) or because migrants to South Africa stay for long periods and therefore were not traceable for this study. By representing men with more local mobility, this study describes a common form of mobility underrecognized in studies on migration and in health and HIV research.

## 5. Conclusions

Contrary to the view that men have full autonomy and avoid HIV services, mobile MLHIV in Malawi experienced high levels of vulnerability and actively tried to stay on ART, even during unpredictable and demanding travel. Mobility was very common, enabling men living in extreme poverty to find work and protect their households against financial catastrophe. However, rigid health systems created inevitable conflict with remaining on ART, especially when travel was unplanned and/or travel duration was longer than expected. As mobility is likely to increase in coming years, HIV programs must adapt to meet the needs of vulnerable, mobile populations.

## Data Availability

Due to the sensitive nature of the data collected and the risk of unwanted disclosure of HIV status of participants, the data in this study will not be made routinely or publicly available. Some data produced in the study may be available upon request to the authors.

## Authors’ contributions

MT and KD conceptualized the study design. MB conducted qualitative data collection; MM and JH supervised quantitative data collection. MT and TT coded qualitative data, and MT, MB, TT, MM, JH, and KD participated in qualitative data analysis. MT and AC conducted quantitative data analysis. MT drafted the initial manuscript. All authors reviewed the manuscript and supported with edits.

## Acknowledgements

The authors would like to acknowledge the participants in the IDEaL and ENGAGE trials, especially the 32 who participated in in-depth interviews, for sharing their experiences and insights with the study team. The authors are also grateful to the research assistants, staff of Partners in Hope (especially Isabella Robson, Khumbo Phiri, Dr. Sam Phiri, and Dr. Joep van Oosterhout), and other mentors (especially Dr. Sundeep Gupta) who supported this endeavor.

## Additional files

Additional file 1: Appendix A

Table showing socio-demographic characteristics and ART history of IDI participants.

## List of abbreviations

ART: antiretroviral therapy
ENGAGE: parent trial
HCW: healthcare worker
IDEaL: parent trial
IDI: in-depth interview
MLHIV: men living with HIV
PLHIV: people living with HIV
MMD: multi-month dispensing
SSA: sub-Saharan Africa

## Appendix A: Socio-demographics and ART history of IDI participants (n=32)

|  |  |
| --- | --- |
| **Demographics** |  |
| Age, median years (IQR) | 39 (34-45) |
| Married, n (%) | 27 (84%) |
| **ART experience** |  |
| Years since diagnosis, median (IQR) | 1.7 (0.8-9.0) |
| Ever initiated ART, n (%) | 30 (94%) |
| Disclosed HIV status to primary partner | 26 of 27 with partners |
| **Socioeconomic status** |  |
| Household access to a cell phone, n (%) | 16 (50%) |
| Has savings, n (%) | 6 (19%) |
| Attended any secondary school, n (%) | 13 (41%) |

